# Association between circadian physical activity patterns and mortality in the UK Biobank

**DOI:** 10.1101/2022.12.05.22283101

**Authors:** Michael J. Stein, Hansjörg Baurecht, Anja M. Sedlmeier, Julian Konzok, Patricia Bohmann, Emma Fontvieille, Laia Peruchet-Noray, Jack Bowden, Christine M. Friedenreich, Béatrice Fervers, Pietro Ferrari, Marc J. Gunter, Heinz Freisling, Michael F. Leitzmann, Vivian Viallon, Andrea Weber

**Affiliations:** Department of Epidemiology and Preventive Medicine, University of Regensburg, Regensburg, Germany; Nutrition and Metabolism Branch, International Agency for Research on Cancer (IARC/WHO), Lyon, France; University of Exeter Medical School, Exeter, U.K.; Department of Cancer Epidemiology and Prevention Research, Cancer Care Alberta, Alberta Health Services, Calgary, AB, Canada; Department of Prevention Cancer Environment, Centre Léon Bérard, Lyon, France; Department of Clinical Sciences, Faculty of Medicine, University of Barcelona, Barcelona, Spain; Novo Nordisk Research Center Oxford, U.K.; Departments of Oncology and Community Health Sciences, Cumming School of Medicine, University of Calgary, Calgary, Alberta, Canada; INSERM UMR1296 Radiation: Defense, Health, Environment, Lyon, France

## Abstract

**Importance:** The benefit of physical activity (PA) for increasing longevity is well-established, however, the impact of diurnal timing of PA on mortality remains poorly understood.

**Objective:** To derive PA patterns and investigate their associations with all-cause mortality.

**Design, Setting, and Participants:** This population-based prospective cohort study analyzed UK Biobank baseline data collected between 2006 and 2010 from adults aged 40 to 79 years in England, Scotland, and Wales. Participants were invited by email to participate in an additional accelerometer study from 2013 to 2015, 7 years (median) after baseline. Participants’ vital status was assessed via linkage with mortality registries through September 2021 (England/Wales) and October 2021 (Scotland). Data analyses were performed in July 2022.

**Exposure:** Loading scores of functional principal components (fPCs) obtained from wrist accelerometer-measured activity metrics. The ‘Euclidean norm minus one’ was used as a summary metric of bodily acceleration aggregated to 24 hourly averages across seven days. These timeseries were used for functional principal component analysis (fPCA).

**Main Outcomes and Measures:** Examination of time-dependent PA patterns obtained using functional principal component analysis in relation to all-cause mortality estimated by multivariable Cox proportional hazard models.

**Results:** Among 96,361 participants (56% female), 2,849 deaths occurred during 6.9 (SD 0.9) years of follow-up. Four distinct functional principal components (fPCs) accounted for 96% of the variation of the accelerometry data. The association of fPC1 and mortality was non-linear (p<0.001). Using a loading score of zero as the reference, a fPC1 score of +2 (high overall PA) was associated with lower mortality (0.91; 95% CI: 0.84–0.99), whereas a score of +1 showed no relation (0.94; 95% CI: 0.89–1.00). A fPC1 score of -2 (low overall PA) was associated with higher mortality (1.71; 95% CI: 1.58–1.84), as was a score of -1 (1.20; 95% CI: 1.13–1.26). A 1-unit score increase on fPC2 (high early day PA) was not associated with mortality (0.97; 95% CI: 0.93–1.02). For fPC3, a 1-unit score increase (high midday PA) was associated with decreased mortality (0.88; 95% CI: 0.84–0.94). In contrast, a 1-unit score increase on fPC4 (high midday and nocturnal PA) was associated with higher mortality (1.14; 95% CI: 1.06– 1.24).

**Conclusions and Relevance:** Higher risks of death were found for patterns denoting lower overall PA and higher late day and nocturnal PA. Conversely, higher levels of PA, distributed continuously, in one, or in two activity peaks during daytime, were inversely associated with lower mortality. Daily timing of PA may have public health implications, as our results suggest that some level of elevated PA during the day and a nighttime rest is associated with longevity.

## 1. Introduction

Physical inactivity is a global concern, with 28% of the world’s population not attaining physical activity (PA) recommendations (1). Such data are disconcerting because of robust evidence on the association of insufficient PA with increased premature mortality (2, 3).

PA is a complex construct, the valid measurement of which is challenging (4). Most evidence on the PA and mortality relation stems from studies using PA self-report methods. Such methods provide data on PA type, dose, and timing but they suffer from PA measurement error (2). Accelerometers are increasingly preferred as PA measure (5) because they show high validity (6) and quantify temporal nuances in movement behaviors (7). However, raw accelerometry data show considerable within and between subject heterogeneity and the data volume and complexity are challenging to analyze (8). Thus, most previous studies have relied on summary accelerometry output instead of comprehensively examining movement and rest profiles throughout the day (9).

Functional principal component analysis (fPCA), an extension of common PCA, is well-suited to analyze temporal patterns and thus represents a promising technique to identify circadian PA patterns. Most previous studies using fPCA have been limited by relying on device-specific summary metrics (‘activity counts’) (10-12) or small sample sizes (10, 12, 13). Few studies have overcome these limitations and those that did showed that fPCs denoting lower or evening PA were positively associated with mortality among older men (14) and that fPCs were associated with socio-demographic characteristics and self-rated health (9).

In this study we aimed to go beyond what has been conducted in previous investigations since it is the first to examine fPCA-based PA patterns in relation to all-cause mortality in a large cohort of men and women.

## 2. Methods

### Study population and data collection

The UK Biobank (UKB) is a prospective cohort study of >500,000 U.K. participants aged 40-69 years when recruited between 2006–2010. The study collected data on sociodemographic and lifestyle factors, and extensive phenotypic information. The assessment visit included a touchscreen questionnaire, interviews, physical and functional measurements, and the collection of biologic samples. The UKB obtained ethics approval from the North West Multi-Centre Research Ethics Committee. All participants provided written informed consent (15).

### Physical activity data

For a subset of 103,679 participants, device-based PA data were available, measured in 2013-2015 using the Axivity AX3 (Newcastle Upon Tyne, UK) wrist-worn triaxial accelerometer for seven days. No participants were pre-excluded from the accelerometer study based on health problems. Participants with valid email addresses were randomly invited to participate in the accelerometer study and were informed that the accelerometer was to be worn continuously on the dominant wrist immediately upon receipt. The device was configured to activate shortly after its arrival and was deactivated seven days later. Subsequently, participants were asked to return the device to the coordinating center. Data processing is detailed elsewhere (16). In brief, the Euclidean norm minus one (ENMO) was derived from raw data. ENMOs represent a summary metric of bodily acceleration measured in milligravity units (m*g*) interpretable as PA volume (Supplement S1). Participants with data from at least 72 hours and data for each one-hour period of the 24-hour cycle (scattered over multiple days) were included, as recommended by the UKB expert working group (N=96,675). Further, we excluded participants with average daily ENMOs above the 99.9 percentile (N=97) and/or missing covariates (N=217), leading to 96,361 participants (=PA pattern sample). We used the average hourly acceleration over seven days, resulting in a 96,361 × 24 matrix.

### Functional Principal Component Analysis (fPCA)

We used fPCA to reduce the dimensionality of the data and to derive PA patterns while retaining information on between-person variation. fPCA calculates components of time series data (= PA data) on which each participant scores with a certain loading. These fPCs, or eigenfunctions, depict the strongest and most important modes of variability in the PA data (18). The loading score, or eigenvalue, reflects the extent to which a participant’s activity data follows a certain pattern (14). Each participant contributes to each identified pattern, either positively (positive loading) or negatively (negative loading). We used fPCA through conditional expectation (PACE), developed for sparse longitudinal data with only few repeated observations per subject (19).

We modeled ENMOs using linear regression, adjusted for age, sex, body mass index, and study center to obtain PA residuals. These were subsequently standardized and subjected to the fPCA. We used a Gaussian kernel smoother and the default for estimating the bandwidth (5% of the observed time range for the mean function; 10% for the covariance function). We tested the robustness of our results in sensitivity analyses using generalized cross-validation for bandwidth selection in conjunction with alternative kernel smoothings. The Epanechnikov kernel is (compared to Gaussian) a compact kernel (|*x* − *x*_0_| ≤ 1) that minimizes (among all kernel smoothers) the asymptotic mean integrated squared error. The number of relevant components was determined using the elbow method, an explained total variability threshold of >95%, and visual inspection of the eigenfunctions (20).

We used the R package *fdapace* to apply the fPCA (21).

### Cohort follow-up and ascertainment of mortality cases

Participants’ vital status was determined through linkage with routine health care data and national death registries (22). Follow-up began at the baseline accelerometry measurement (June 2013 to December 2015) and ended at the date of complete follow-up (September 2021 for England/Wales or October 2021 for Scotland) (23), lost-to-follow-up, or date of death. All-cause mortality was considered the endpoint.

### Covariates

We identified potential confounding covariables *a priori* based on evidence-derived directed acyclic graphs (24) (Supplement S2). The main model was stratified by sex and study center and was further adjusted for self-reported prevalent diabetes, prevalent cardiovascular disease (CVD; combination of heart attack, angina pectoris, stroke, and high blood pressure), smoking status (never; former; current), alcohol consumption status (never; former; current), socio-economic status (Townsend Index of Deprivation), education level (college or university degree; A levels/AS levels or equivalent, NVQ or HND or HNC or equivalent, other professional qualifications; O levels/GCSEs or equivalent, CSEs or equivalent; none of the above), sedentary behavior (sum of time spent watching TV, using a computer during leisure time, and driving for transportation), and diet (healthy diet score (25) adjusted to a 0–7 scale). These covariates were measured at baseline (2006–2010), i.e., before accelerometry assessment.

### Statistical analysis

Statistical analysis was conducted with complete data after removing missing covariate data (Supplement S3). Participants with prevalent cancer (pre accelerometry; cancer registry data) (N=13,270) were excluded to minimize reverse causation (26). Ultimately, we included 81,765 participants in our assessment of all-cause mortality (=mortality sample).

Cox proportional hazards regression with age as the underlying time metric (27) was used to estimate hazard ratios (HR) and corresponding 95% confidence intervals (CI) for associations between PA patterns and mortality. Non-linearity was accounted for by restricted cubic splines with four knots at the 0.05, 0.35, 0.65, and 0.95 quantiles. Departure from linearity was tested for all variables by testing the coefficient of the second and third spline transformation equal to zero (28). Proportional hazard assumptions were checked using Schoenfeld residuals and visually. We conducted several sensitivity analyses to evaluate the robustness of our results. Specifically, we excluded deaths that occurred within two years after accelerometry assessment; excluded participants with prevalent comorbidities; and used smoking intensity (pack years) and alcohol use intensity (grams per day) as covariates (29). We examined interaction by sedentary behavior and age. To investigate the robustness of our derived PA patterns, we tested different fPCA hyperparameters (kernel smoother and bandwidths).

Cox regression was conducted using the *rms* package (30). All data processing and statistical analyses were performed using R 4.2.0 (31).

## 3. Results

We obtained PA patterns from 96,361 participants (56.3% female). Participants were 61.9±7.9 years old at accelerometry assessment. The median follow-up time was 6.9±0.9 years, during which 2,849 participants (3.0%) died. The average daily ENMO was 28.0±8.4 m*g*. There was no meaningful deviation for those who were excluded due to missing covariate data (27.0±8.4 m*g*). Baseline characteristics for excluded participants did not differ from the population for analysis (Supplement S4).

### Physical activity patterns

The fPCA revealed four fPCs that accounted for 95.8% of the total variance in the temporal distribution of PA during the day (Figure 1A). No clear elbow was visible and further eigenfunctions showed no patterns that provided interpretable added value. The first fPC (fPC1) explained 65.5% of the variability denoting overall PA during the day. The second component fPC2 accounted for 17.0% of the variance depicting the contrast between early and late hours. The third component fPC3 explained 9.0% of the variance depicting the contrast of midday and early/late hours. A similar pattern was found for fPC4 (4.3% explained variance), except that fPC4 represented the contrast of midday/night and morning/evening hours.

**Figure 1.**
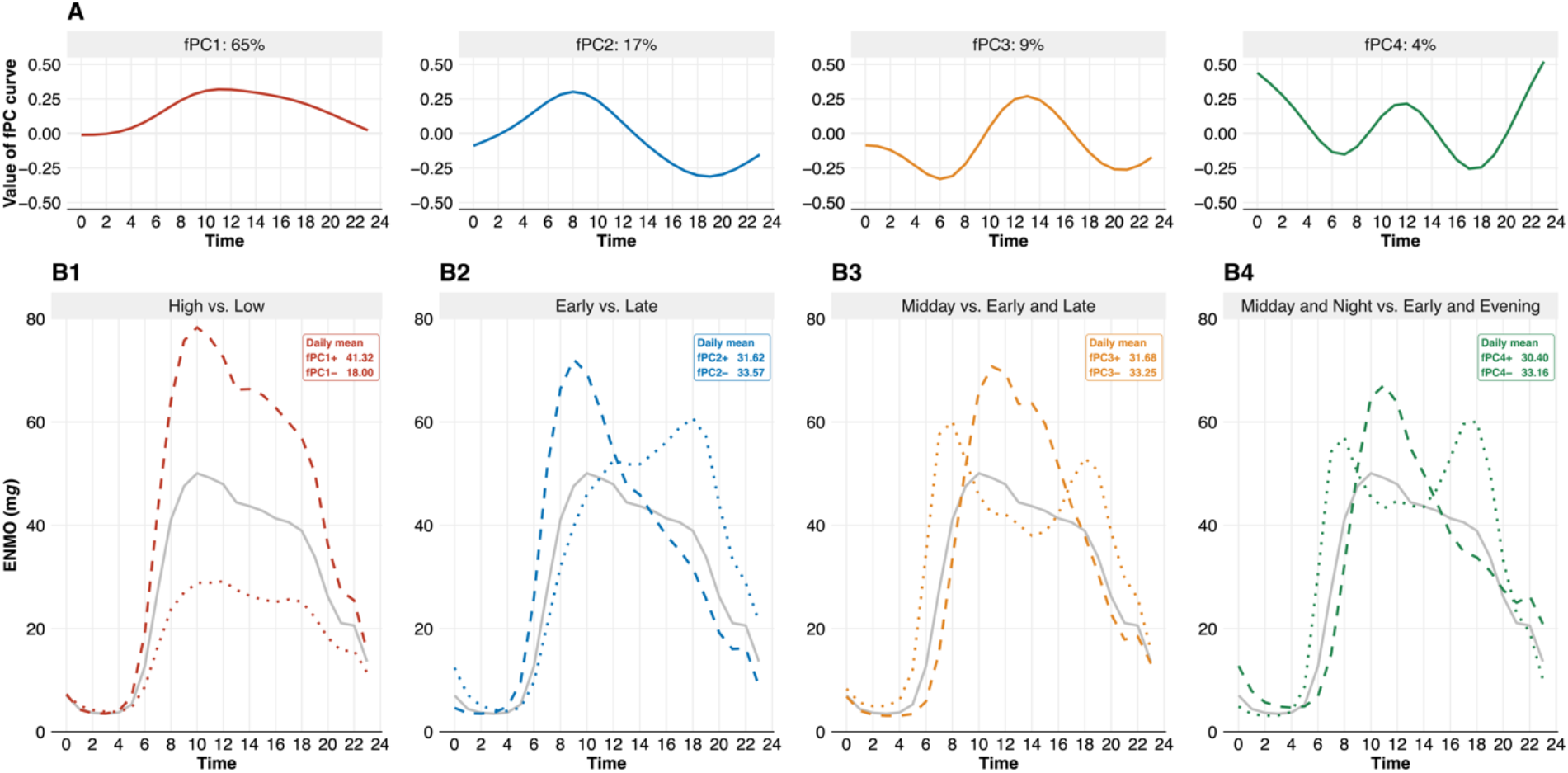
A: The first four eigenfunctions. B: The average daily time course of PA. The solid grey line represents the population PA average (ENMOs in mg), the dashed line represents the positive scorers (at least one standard deviation away from the mean score), and the dotted line represents the negative scorers (at least one standard deviation away). Daily average ENMOs were similar for positive and negative scores on fPC2, fPC3, and fPC4 as well as between these fPCs.

For better interpretability, we focused on participants who scored positive (≥1 SD above the mean) or negative (≤1 SD below the mean) on a given component (Table 2). High and low values of the fPC curve (large weights) correspond to increased PA levels during these time periods (Figure 1B). Positive scores on fPC1 were related to increased PA levels between 6AM-10PM. Positive scores on fPC2 showed early PA (8AM-12PM); on fPC3 midday PA (10AM-4PM); and on fPC4 midday PA (10AM-4PM) and nighttime PA (12AM-4AM). Negative scores were related to inverse patterns (Table 2).

**Table 1.** Descriptive baseline characteristics.

| Variable | PA pattern sample<br>(N = 96,361) | Mortality sample<br>(N = 81,765) |
| --- | --- | --- |
| Sex (%) |  |  |
| *Female* | 54,279 (56.3) | 45,502 (55.7) |
| *Male* | 42,082 (43.7) | 36,259 (44.4) |
| Age at baseline (sd) | 56.16 (7.82) | 55.68 (7.83) |
| Age at accelerometry (sd) | 61.85 (7.85) | 61.34 (7.85) |
| Age at exit (sd) | 68.67 (7.78) | 68.19 (7.79) |
| Body mass index (sd) | 26.71 (4.53) | 26.71 (4.53) |
| Diet score (sd) | 3.71 (1.33) | 3.70 (1.33) |
| Socio-economic status (sd) | -1.73 (2.82) | -1.70 (2.83) |
| Sedentary behavior (sd) | 4.29 (2.46) | 4.29 (2.47) |
| Smoking status (%) |  |  |
| *Never* | 54,878 (57.0) | 47,160 (57.7) |
| *Former* | 34,578 (35.0) | 28,914 (35.4) |
| *Current* | 6,660 (6.9) | 5,687 (7.0) |
| Pack years of smoking (sd) | 20.25 (16.66) | 6.49 (13.24) |
| Alcohol drinking status (%) |  |  |
| *Never* | 2,790 (2.9) | 2,386 (2.9) |
| *Former* | 2,655 (2.8) | 2,254 (2.8) |
| *Current* | 90,839 (94.3) | 77,121 (94.3) |
| Alcohol intake in grams/day (sd) | 16 (17) | 16 (17) |
| Qualifications (%) |  |  |
| *College or university degree* | 41,413 (43.0) | 35,848 (43.9) |
| *A levels/AS levels or equivalent,* |  |  |
| *NVQ or HND or HNC or equivalent,* |  |  |
| *Other professional qualifications* | 22,603 (23.5) | 19,371 (23.7) |
| *O levels/GCSEs or equivalent, CSEs* |  |  |
| *or equivalent* | 23,419 (24.3) | 20,031 (24.5) |
| *None of the above* | 7,973 (8.3) | 6,511 (8.0) |
| Cancer (%) |  |  |
| *Before accelerometry* | 13,270 (13.8) | *excluded* |
| *No* | 76,412 (79.3) | 75,188 (92.0) |
| *After baseline* | 6,679 (6.9) | 6,577 (8.0) |
| Diabetes (%) |  |  |
| *No* | 92,860 (96.4) | 79,004 (96.6) |
| *Yes* | 3,328 (3.5) | 2,757 (3.4) |
| Cardiovascular disease (%) |  |  |
| *No* | 72,196 (74.9) | 61,930 (75.8) |
| *Yes* | 24,035 (24.9) | 19,831 (24.3) |

The PA pattern sample consists of all participants with valid accelerometry data (those with missing body mass index were excluded). For the mortality sample, we excluded subjects with pre-baseline cancer and those with missing covariates.

**Table 2.**
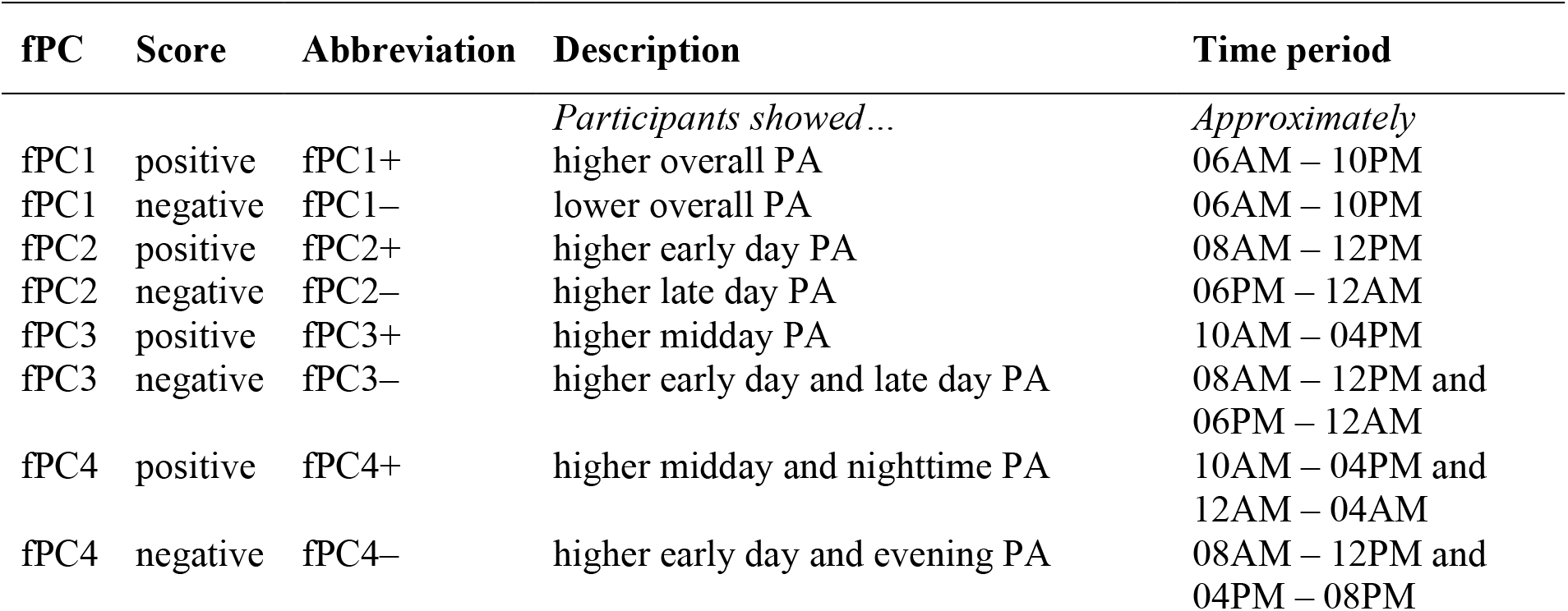
Description of PA patterns.

Sensitivity analyses showed that results were robust for changes in the parameters used to determine fPCs. Alternatively, when an Epanechnikov kernel for smoothing was used, the explained variability was smaller for fPC1 (∼16% decrease) but higher for fPC3 and fPC4, and more than four components were necessary to reach the 95% threshold (Supplement S5A). Nevertheless, the shape of the first four eigenfunctions remained similar (Supplement S5B). Variation in the bandwidths of the kernel smoothers did not affect the explained variance.

### Mortality

We noted a strong non-linear relation of fPC1 to mortality (non-linear term p < 0.001). Negative scores (lower overall PA) were associated with increased mortality. With a loading score of zero as the reference, fPC1 scores of -2 and -1 were related to elevated mortality, with HRs of 1.71 (95% CI: 1.58–1.84, p < 0.001) and 1.20 (95% CI: 1.13–1.26, p < 0.001), respectively.

Conversely, scoring +2 (higher overall PA) was associated with reduced mortality (0.91; 95% CI: 0.84–0.99, p < 0.001). We found no association between fPC2 (higher early day PA) and mortality (0.97; 95% CI: 0.93–1.02, p=0.26). fPC3 (higher midday PA) was associated with decreased mortality – for a one unit score increase, the HR was 0.88 (95% CI: 0.84–0.94, p < 0.001). An association in the opposite direction was apparent for fPC4, i.e., higher midday and nighttime PA (1.14; 95% CI: 1.06–1.24, p < 0.001). Figure 2A displays the HRs for specific fPC loading scores in relation to the reference score zero. Figure 2B shows the (non-)linear relation of the component scores to all-cause mortality.

**Figure 2.**
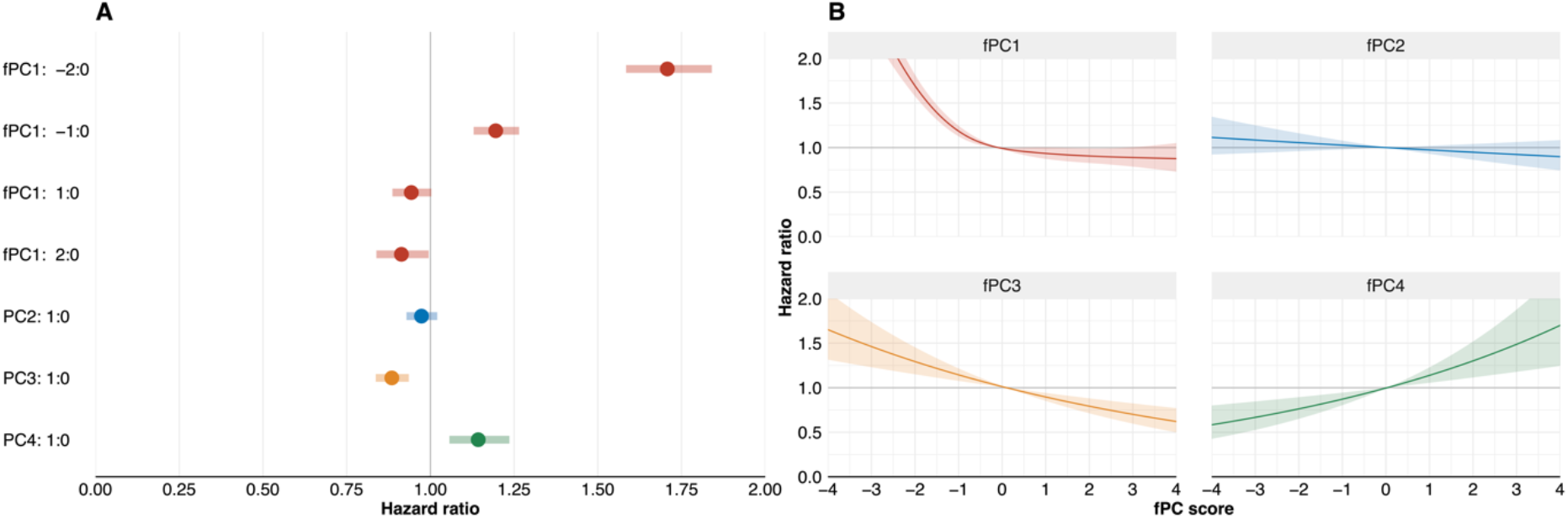
A: HRs for all four components for the scores of -2, -1, 1, and 2 with reference to a score of 0. B: (Non-)linear relation of continuous fPC scores and HRs.

In addition, in a sensitivity model we examined the impact of smoking intensity (pack years) and alcohol intensity (grams per day) and found no deviation from our main results (Table 3). The associations also remained apparent when using another kernel smoother, with non-significant estimates for fPC4 (Supplement S5C). In an additional model, we excluded deaths within two years after accelerometry assessment (N=336) and it appeared, that fPC4 was no longer related to mortality (1.08; 95% CI: 0.99–1.18, p=0.875). The exclusion of participants with CVD and/or diabetes at baseline (N=20,729) led to wider CIs for positive scores on fPC1, which now included the null value, but the effect direction remained compared to the main model (0.91; 95% CI: 0.81–1.02) (Supplements S6). Lastly, we examined potential interactions between the fPCs scores and sedentary behavior and age. None of the interaction terms were statistically significant except for the non-linear modelling of fPC1 (p=0.04), where high positive scores for the age group of 60-69 years showed decreased risks (score of +3: 0.81; 95% CI: 0.68–0.98). However, the association direction for all other age groups was reversed for positive scores on fPC1, with very wide confidence intervals, all containing the null value (Supplement S7).

**Table 3.** Hazard ratios of the four principal components for all-cause mortality.

| Component | Main model (N=81,765)<br>HR (95% CI) | Sensitivity model (N=60,446)<br>HR (95% CI) |
| --- | --- | --- |
| fPC1 |  |  |
| -2:0 | 1.71 (1.59–1.84) | 1.77 (1.62–1.93) |
| -1:0 | 1.20 (1.13–1.27) | 1.22 (1.14–1.30) |
| 1:0 | 0.94 (0.88–1.00) | 0.93 (0.87–1.00) |
| 2:0 | 0.91 (0.84–0.99) | 0.89 (0.81–0.99) |
| Overall *p*-value | $1.61 \times 10^{-59}$ | $1.96 \times 10^{-49}$ |
| fPC2 | 0.97 (0.93–1.02) | 0.98 (0.93–1.04) |
| Overall *p*-value | 0.274 | 0.550 |
| fPC3 | 0.89 (0.84–0.94) | 0.89 (0.83–0.95) |
| Overall *p*-value | $2.64 \times 10^{-5}$ | $6.97 \times 10^{-4}$ |
| fPC4 | 1.14 (1.05–1.23) | 1.12 (1.02–1.23) |
| Overall *p*-value | $1.18 \times 10^{-3}$ | $2.16 \times 10^{-2}$ |

HRs are reported for a one score increase except for fPC1, where HRs for a score of -2, -1, 1, and 2 compared to 0 are reported. The main model was adjusted for smoking status and alcohol status. The sensitivity model was adjusted for pack years of smoking and alcohol grams per day.

## 4. Discussion

We derived novel circadian PA patterns using fPCA and uncovered four eigenfunctions that explained almost the entire variability of 24h-accelerometry data in the UK Biobank. These patterns described the time course of activity throughout the day and differences between morning and evening hours. We found that three fPCs were associated with mortality. Positive fPC1 scores representing higher overall PA showed decreased mortality, whereas negative fPC1 scores reflecting lower overall PA displayed higher mortality. Negative scores on fPC3, signifying the combination of higher early day and late day PA were related to increased mortality, whereas positive fPC3 scores representing higher midday PA were associated with lower mortality. Moreover, positive fPC4 scores capturing higher midday and nighttime PA were associated with higher mortality, whereas negative fPC4 scores symbolizing the combination of higher early day and evening time PA were related to lower mortality. By comparison, fPC2 was unrelated to mortality.

Our results point towards the importance of overall PA for longevity. Inadequate levels of PA increase the risk of premature mortality, whereas high PA levels are associated with lower risk of early death (32, 33). Our findings regarding fPC1 support this evidence. Specifically, lower fPC1 scores were associated with greater mortality hazard, and scoring highly positive on fPC1 was associated with lower premature mortality. The inverse relation of fPC1 to mortality was attenuated above a score of 2, which mirrors the World Health Organization (WHO) guidelines on PA and sedentary behavior stating that the beneficial effects of activity diminish at higher PA levels (34). In addition, interaction by age indicated that the effects of positive fPC1 scores might depend on age.

Moreover, our patterns revealed insights into the time course of activity that go beyond the general time course of PA. Positive fPC3 scores (fPC3+; one peak at midday) and negative fPC4 scores (fPC4–; two peaks, morning and evening) showed above-average activity levels and were related to decreased mortality. While analyses of biological response to exercise proposed that circadian timing of PA may matter (35, 36), our results on mortality suggest that it is not the shape of the activity that matters if a certain minimum level of (above-average) PA is achieved. In addition, our results indicate that nighttime activity is adversely related to longevity, consistent with observations that later sleep timing is associated with adverse health effects and mortality (37, 38). Both fPC3– and particularly fPC4+ showed increased activity levels during night hours, a pattern associated with higher mortality. Increased nighttime activity might be confounded by occupation (39), lifestyle factors (38), or genetics (40). Further, subclinical diseases might cause sleep disruptions which could explain the slightly increased activity levels at night suggesting potential reverse causation. When we excluded deaths within the first two years after the PA assessment, fPC4 was no longer statistically significantly associated with mortality. Of note, a recent study applied k-means clustering to UKB accelerometer data and found that nighttime activity was associated with increased CVD incidence compared to morning PA (41).

We present novel analyses of PA data using the residuals of raw accelerometry-based PA metrics. Nevertheless, the shapes of our PA patterns are similar to previous applications of fPCA to accelerometry data (9-11, 13, 14), which provides confidence in the robustness of our results. One study derived four patterns (88% variance explained) among 2,976 men and found that the first component denoted overall activity, with lower quartiles showing higher mortality hazards (14). Another study derived four fPCs (87% variance explained) and reported associations with population characteristics and self-reported health among 7,657 individuals (9). We greatly expand on these findings by presenting four fPCs explaining 96% of the variability of the data on 96,361 participants. Moreover, we used ENMOs and thus overcame the limitations of summary counts. We also used different kernel smoothers and bandwidths in sensitivity analyses to provide more robust results. Finally, we did not use the accelerometer data directly as input for the fPCA, but instead used the residuals, which were *a priori* adjusted for potential major confounders.

Our study has some limitations. First, at least two years elapsed between measurement of covariates at baseline and accelerometer measurement. During this time, important changes may have occurred. Covariates might have changed, or the participants may have developed comorbidities. Additionally, it is possible that participants altered their behavior because they were aware of wearing an accelerometer (‘reactivity’) (42). Differential misclassification of the exposure could lead to biased associations with the outcome. Ultimately, the general limitations of accelerometry remain, including lack of data on context, which limits the interpretation and comparability of our results with context-based measurements.

Notwithstanding these limitations, we were able to gain novel insights. The valuable information of the temporal distribution of PA is underutilized, with only few studies examining PA patterns (43). We addressed this knowledge gap by conducting a robust study of PA patterns based on a large sample and associations with mortality. By using an unsupervised approach, we made no *a priori* assumptions. Our patterns were robust to different fPCA settings (smoothing kernel, varying bandwidths) and covariate modelling. Compared with previous research, we examined a significantly larger sample as well as a longer time period, and hence, presented stable effect estimates with smaller CIs. While previous accelerometer studies were based on summary statistics, we used a metric derived from raw data, which facilitates comparability and interpretability.

## 5. Conclusion

Our study addressed a gap in previous literature regarding the temporal course of PA and its association with mortality. We found novel circadian patterns of PA using fPCA. These patterns were defined as the time course of PA over 24 hours. Four eigenfunctions explained most of the variation in the data and the patterns were related to all-cause mortality. Our results indicate that it is less important during which daytime hours one is active but rather, engaging in a minimum level of PA and resting during the late evening and nighttime hours is associated with decreased mortality. Future studies need to confirm the validity and robustness of our methods and results. Finally, contextual information such as activity type would be of additional value.

## Supporting information

STROBE checklist

## Data Availability

Bona fide researchers can apply to use the UK Biobank dataset by registering and applying at http://ukbiobank.ac.uk/register-apply/.

## Acknowledgements

UK Biobank is an open access resource. Bona fide researchers can apply to use the UK Biobank dataset by registering and applying at http://ukbiobank.ac.uk/register-apply/.

This research has been conducted using the UK Biobank Resource under Application Number 55870 and we express our gratitude to the participants and those involved in building the resource.

## Literature

1. Guthold R, Stevens GA, Riley LM, Bull FC. Worldwide trends in insufficient physical activity from 2001 to 2016: a pooled analysis of 358 population-based surveys with 1.9 million participants. The Lancet Global Health. 2018;6(10):e1077–e86.

2. Arem H, Moore SC, Patel A, Hartge P, Berrington de Gonzalez A, Visvanathan K, et al. Leisure Time Physical Activity and Mortality: A Detailed Pooled Analysis of the Dose-Response Relationship. JAMA Internal Medicine. 2015;175(6):959–67.

3. McKinney J, Lithwick DJ, Morrison BN, Nazarri H, Isserow S, Heilbron B, et al. The health benefits of physical activity and cardiorespiratory fitness. BCMJ. 2016;58(3):131–7.

4. Leitzmann M, Powers H, Anderson AS, Scoccianti C, Berrino F, Boutron-Ruault M-C, et al. European Code against Cancer 4th Edition: Physical activity and cancer. Cancer Epidemiology. 2015;39:S46–S55.

5. Gao Z, Liu W, McDonough DJ, Zeng N, Lee JE. The Dilemma of Analyzing Physical Activity and Sedentary Behavior with Wrist Accelerometer Data: Challenges and Opportunities. Journal of Clinical Medicine. 2021;10(24):5951.

6. Sallis JF, Saelens BE. Assessment of physical activity by self-report: status, limitations, and future directions. Research Quarterly for Exercise and Sport. 2000;71 Suppl 2:1–14.

7. Clark S, Lomax N, Morris M, Pontin F, Birkin M. Clustering Accelerometer Activity Patterns from the UK Biobank Cohort. Sensors. 2021;21(24):8220.

8. Karas M, Bai J, Strączkiewicz M, Harezlak J, Glynn NW, Harris T, et al. Accelerometry Data in Health Research: Challenges and Opportunities. Stat Biosci. 2019;11(2):210–37.

9. Xiao Q, Lu J, Zeitzer JM, Matthews CE, Saint-Maurice PF, Bauer C. Rest-activity profiles among U.S. adults in a nationally representative sample: a functional principal component analysis. International Journal of Behavioral Nutrition and Physical Activity. 2022;19(1):32.

10. Gershon A, Ram N, Johnson SL, Harvey AG, Zeitzer JM. Daily Actigraphy Profiles Distinguish Depressive and Interepisode States in Bipolar Disorder. Clinical Psychological Science. 2016;4(4):641–50.

11. Leroux A, Di J, Smirnova E, McGuffey EJ, Cao Q, Bayatmokhtari E, et al. Organizing and analyzing the activity data in NHANES. Stat Biosci. 2019;11(2):262–87.

12. Xu SY, Nelson S, Kerr J, Godbole S, Johnson E, Patterson RE, et al. Modeling Temporal Variation in Physical Activity Using Functional Principal Components Analysis. Stat Biosci. 2019;11(2):403–21.

13. Difrancesco S, Riese H, Merikangas KR, Shou H, Zipunnikov V, Antypa N, et al. Sociodemographic, Health and Lifestyle, Sampling, and Mental Health Determinants of 24-Hour Motor Activity Patterns: Observational Study. J Med Internet Res. 2021;23(2):e20700.

14. Zeitzer JM, Blackwell T, Hoffman AR, Cummings S, Ancoli-Israel S, Stone K, et al. Daily Patterns of Accelerometer Activity Predict Changes in Sleep, Cognition, and Mortality in Older Men. The Journals of Gerontology: Series A. 2017;73(5):682–7.

15. Sudlow C, Gallacher J, Allen N, Beral V, Burton P, Danesh J, et al. UK Biobank: An Open Access Resource for Identifying the Causes of a Wide Range of Complex Diseases of Middle and Old Age. PLOS Medicine. 2015;12(3):e1001779.

16. Doherty A, Jackson D, Hammerla N, Plötz T, Olivier P, Granat MH, et al. Large Scale Population Assessment of Physical Activity Using Wrist Worn Accelerometers: The UK Biobank Study. PLOS ONE. 2017;12(2):e0169649.

17. Ramakrishnan R, Doherty A, Smith-Byrne K, Rahimi K, Bennett D, Woodward M, et al. Accelerometer measured physical activity and the incidence of cardiovascular disease: Evidence from the UK Biobank cohort study. PLOS Medicine. 2021;18(1):e1003487.

18. Ramsay JO, Silverman BW. Functional Data Analysis. 2 ed: Springer New York, NY; 2005.

19. Yao F, Müller H-G, Wang J-L. Functional Data Analysis for Sparse Longitudinal Data. ournal of the American Statistical Association. 2005;100(470):577–90.

20. James G, Witten D, Hastie T, Tibshirani R. An Introduction to Statistical Learning: with Applications in R. 2 ed. New York, NY: Springer; 2021.

21. Gajardo A, Bhattacharjee S, Carroll C, Chen Y, Dai X, Fan J, et al. fdapace: Functional Data Analysis and Empirical Dynamics. R package version 0.5.8. 2021 [Available from: https://CRAN.R-project.org/package=fdapace.

22. Trehearne A. Genetics, lifestyle and environment. Bundesgesundheitsblatt - Gesundheitsforschung - Gesundheitsschutz. 2016;59(3):361–7.

23. UK Biobank. Data providers and dates of data availability 2022 [Available from: https://biobank.ndph.ox.ac.uk/showcase/exinfo.cgi?src=Data_providers_and_dates.

24. VanderWeele TJ. Principles of confounder selection. European Journal of Epidemiology. 2019;34(3):211–9.

25. Lourida I, Hannon E, Littlejohns TJ, Langa KM, Hyppönen E, Kuźma E, et al. Association of Lifestyle and Genetic Risk With Incidence of Dementia. JAMA. 2019;322(5):430–7.

26. Strain T, Wijndaele K, Sharp SJ, Dempsey PC, Wareham N, Brage S. Impact of follow-up time and analytical approaches to account for reverse causality on the association between physical activity and health outcomes in UK Biobank. International Journal of Epidemiology. 2019;49(1):162–72.

27. Cologne J, Hsu W-L, Abbott RD, Ohishi W, Grant EJ, Fujiwara S, et al. Proportional Hazards Regression in Epidemiologic Follow-up Studies: An Intuitive Consideration of Primary Time Scale. Epidemiology. 2012;23(4):565–73.

28. Harrell FE. Regression Modeling Strategies. With Applications to Linear Models, Logistic and Ordinal Regression, and Survival Analysis: Springer Cham; 2015.

29. Bradbury KE, Murphy N, Key TJ. Diet and colorectal cancer in UK Biobank: a prospective study. International Journal of Epidemiology. 2019;49(1):246–58.

30. Harrell F. rms: Regression Modeling Strategies. R package version 6.3-0 2022 [Available from: https://CRAN.R-project.org/package=rms.

31. R Core Team. R: A language and environment for statistical computing Vienna, Austria: R Foundation for Statistical Computing; 2022 [Available from: https://www.R-project.org/.

32. Biswas A, Oh PI, Faulkner GE, Bajaj RR, Silver MA, Mitchell MS, et al. Sedentary Time and Its Association With Risk for Disease Incidence, Mortality, and Hospitalization in Adults. Annals of Internal Medicine. 2015;162(2):123–32.

33. Ekelund U, Tarp J, Steene-Johannessen J, Hansen BH, Jefferis B, Fagerland MW, et al. Dose-response associations between accelerometry measured physical activity and sedentary time and all cause mortality: systematic review and harmonised meta-analysis. BMJ. 2019;366:4570.

34. Bull FC, Al-Ansari SS, Biddle S, Borodulin K, Buman MP, Cardon G, et al. World Health Organization 2020 guidelines on physical activity and sedentary behaviour. British Journal of Sports Medicine. 2020;54(24):1451–62.

35. Sato S, Dyar KA, Treebak JT, Jepsen SL, Ehrlich AM, Ashcroft SP, et al. Atlas of exercise metabolism reveals time-dependent signatures of metabolic homeostasis. Cell Metabolism. 2022;34(2):329-45.e8.

36. Savikj M, Gabriel BM, Alm PS, Smith J, Caidahl K, Björnholm M, et al. Afternoon exercise is more efficacious than morning exercise at improving blood glucose levels in individuals with type 2 diabetes: a randomised crossover trial. Diabetologia. 2019;62(2):233–7.

37. Knutson KL, von Schantz M. Associations between chronotype, morbidity and mortality in the UK Biobank cohort. Chronobiology International. 2018;35(8):1045–53.

38. Yi J, Wang L, Guo J, Sun P, Shuai P, Ma X, et al. Association of nighttime physical activity with all-cause and cardiovascular mortality: Results from the NHANES. Frontiers in Cardiovascular Medicine. 2022;9.

39. Jørgensen JT, Karlsen S, Stayner L, Hansen J, Andersen ZJ. Shift work and overall and cause-specific mortality in the Danish nurse cohort. Scandinavian Journal of Work, Environment & Health. 2017(2):117–26.

40. Jones SE, van Hees VT, Mazzotti DR, Marques-Vidal P, Sabia S, van der Spek A, et al. Genetic studies of accelerometer-based sleep measures yield new insights into human sleep behaviour. Nature Communications. 2019;10(1):1585.

41. Albalak G, Stijntjes M, van Bodegom D, Jukema JW, Atsma DE, van Heemst D, et al. Setting your clock: associations between timing of objective physical activity and cardiovascular disease risk in the general population. European Journal of Preventive Cardiology. 2022.

42. Jiménez-Buedo M. Reactivity in social scientific experiments: what is it and how is it different (and worse) than a Placebo effect? European Journal for Philosophy of Science. 2021;11(2):42.

43. Niemelä M, Kangas M, Farrahi V, Kiviniemi A, Leinonen A-M, Ahola R, et al. Intensity and temporal patterns of physical activity and cardiovascular disease risk in midlife. Preventive Medicine. 2019;124:33–41.

