## Supplemental material for "Association between circadian physical activity patterns and mortality in the UK Biobank"

Preprint

### Supplemental Materials

S1 The Euclidean norm minus one (ENMO)
S2 Directed Acyclic Graph
S3 Flowchart for participant inclusion
S4 Descriptive baseline characteristics of excluded participants
S5A Sensitivity fPCA with different bandwidth estimations and kernel smoothers
S5B The first four eigenfunctions (A) and positive and negative scorers (B) when using an Epanechnikov kernel
S5C Hazard ratios when using an Epanechnikov kernel
S6 Cox models without deaths within 2 years after accelerometry and without prevalent CVD and/or diabetes
S7 Interaction of fPC1 and age groups

**S1: The Euclidean norm minus one (ENMO)**
ENMOs are the Euclidean norm minus one, as described by van Hees et al. (1). They are defined as the Euclidean norm for the three-dimensional acceleration for each time point with one gravitational unit being subtracted and negative values truncated to zero:

ENMO = $\sqrt{x^2+y^2+z^2}-1g$

In the UKB, ENMOs were collapsed to five-second epoch levels measured in milli gravity (m$g$) units. Hence, for each participant, up to approximately 120,000 ENMOs could be measured over the 7-day period.

1. van Hees VT, Gorzelniak L, Dean León EC, Eder M, Pias M, Taherian S, et al. Separating Movement and Gravity Components in an Acceleration Signal and Implications for the Assessment of Human Daily Physical Activity. PLOS ONE. 2013;8(4):e61691.

Preprint

**S2: Directed Acyclic Graph**

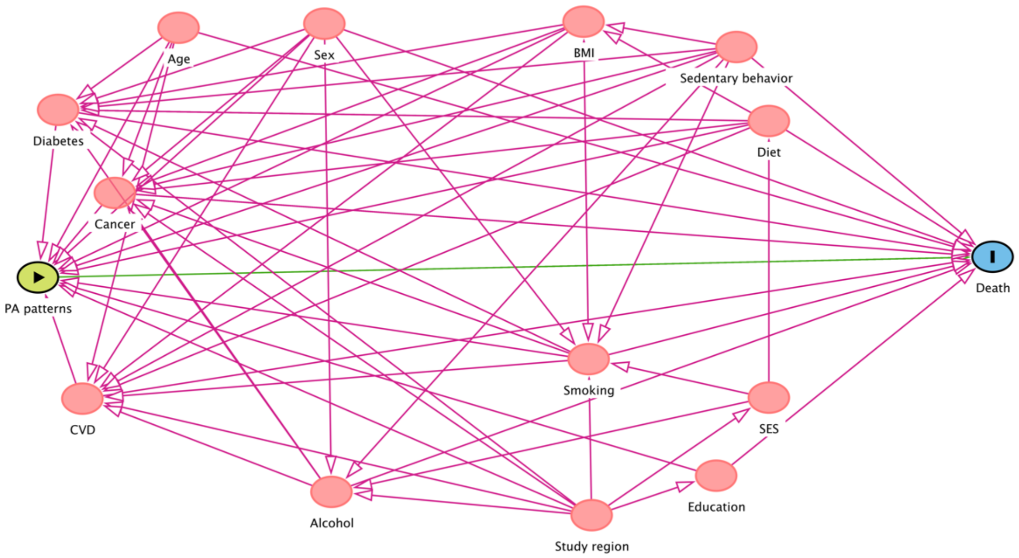


Abbreviations: PA – Physical activity, BMI – Body mass index, CVD – Cardiovascular disease, SES – Socio-economic status.

Preprint

### S3: Flowchart for participant inclusion

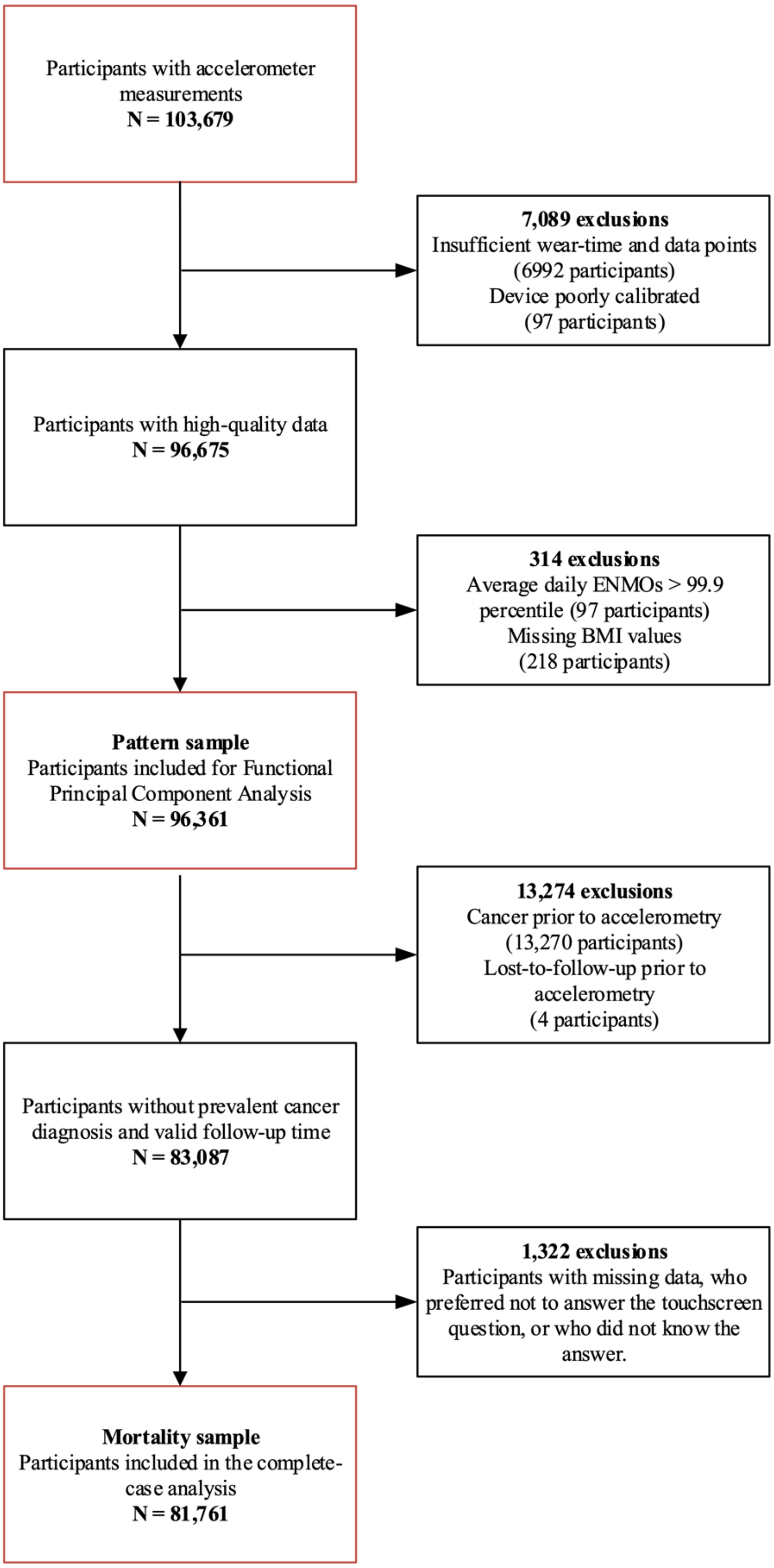

**S4: Descriptive baseline characteristics of excluded participants**

| Variable | Excluded due to poor accelerometry data (N = 6,960) | Excluded due to missing covariate data (N = 1,322) |
|---|---|---|
| Sex (%) | | |
| *Female* | 3,821 (54.9) | 721 (54.54) |
| *Male* | 3,139 (45.10) | 601 (45.46) |
| Age at baseline (sd) | 54.69 (7.99) | 56.08 (8.04) |
| Age at accelerometry (sd) | 60.36 (7.99) | 62.76 (7.99) |
| Age at exit (sd) | 67.14 (7.92) | 69.44 (7.90) |
| Body mass index (sd) | 27.05 (4.77) | 27.05 (4.91) |
| Diet score (sd) | 3.70 (1.32) | 3.62 (1.33) |
| Socio-economic status (sd) | -1.47 (2.96) | -1.32 (3.06) |
| Sedentary behavior (sd) | 4.38 (2.60) | 4.20 (2.68) |
| Smoking status (%) | | |
| *Never* | 3,871 (55.62) | 628 (47.50) |
| *Former* | 2,481 (35.65) | 380 (28.74) |
| *Current* | 591 (8.49) | 100 (7.56) |
| Pack years of smoking (sd) | 19.79 (16.85) | 6.53 (14.76) |
| Alcohol drinking status (%) | | |
| *Never* | 194 (2.79) | 48 (3.63) |
| *Former* | 191 (2.74) | 40 (3.03) |
| *Current* | 6,570 (94.4) | 1,163 (87.97) |
| Alcohol intake in grams/day (sd) | 17.11 (16.96) | 16.72 (18.64) |
| Qualifications (%) | | |
| *College or university degree* | 3,066 (44.05) | 116 (8.77) |
| *A levels/AS levels or equivalent, NVQ or HND or HNC or equivalent, Other professional qualifications* | 1,636 (23.51) | 175 (13.24) |
| *O levels/GCSEs or equivalent, CSEs or equivalent* | 1,726 (24.8) | 115 (8.70) |
| *None of the above* | 461 (6.62) | 106 (8.02) |
| Cancer (%) | | |
| *Before accelerometry* | 900 (12.93) | *excluded* |
| *No* | 5,610 (80.60) | 1,220 (92.28) |
| *After baseline* | 450 (6.47) | 102 (7.72) |
| Diabetes (%) | | |
| *No* | 6,720 (96.55) | 1,119 (84.64) |
| *Yes* | 229 (3.29) | 53 (4.01) |
| Cardiovascular disease (%) | | |
| *No* | 5,362 (77.04) | 846 (63.99) |
| *Yes* | 1,591 (22.86) | 361 (27.31) |

The proportions of participants excluded due to missing covariate data do not necessarily sum up to 100% due to missing data.

Preprint

### S5A: Sensitivity fPCA with different bandwidth estimations and kernel smoothers

| | Fraction of variance explained | | | | | | | | |
|---|---|---|---|---|---|---|---|---|---|
| | *K* | | | | | | | | |
| | 1 | 2 | 3 | 4 | 5 | 6 | 7 | 8 | 9 |
| **Gaussian** | | | | | | | | | |
| Default | 65.49% | 17.01% | 9.00% | 4.32%* | 2.95% | 0.73% | - | - | - |
| GCV | 70.29% | 13.80% | 9.19% | 3.25%* | 2.21% | 0.86% | - | - | - |
| GMeanGCV | 66.29% | 15.57% | 9.37% | 4.16%* | 3.16% | 0.94% | - | - | - |
| **Epanechnikov** | | | | | | | | | |
| Default | 50.33% | 16.62% | 13.82% | 6.74% | 4.78% | 2.97%* | 2.26% | 1.08% | 0.97% |
| GCV | 49.47% | 15.28% | 13.26% | 9.22% | 5.21% | 2.84%* | 2.15% | 1.16% | 0.94% |
| GMeanGCV | 49.80% | 16.30% | 13.58% | 7.22% | 5.29% | 2.98%* | 2.25% | 1.10% | 1.02% |

*Cumulative fraction of variance explained above 95%.

Note: Default refers to the default settings of the fPCA function in fdapace. GCV is Generalized Cross-Validation; GMeanGCV is Geometric Mean and GCV.

### S5B: The first four eigenfunctions (A) and positive and negative scorers (B) when using an Epanechnikov kernel

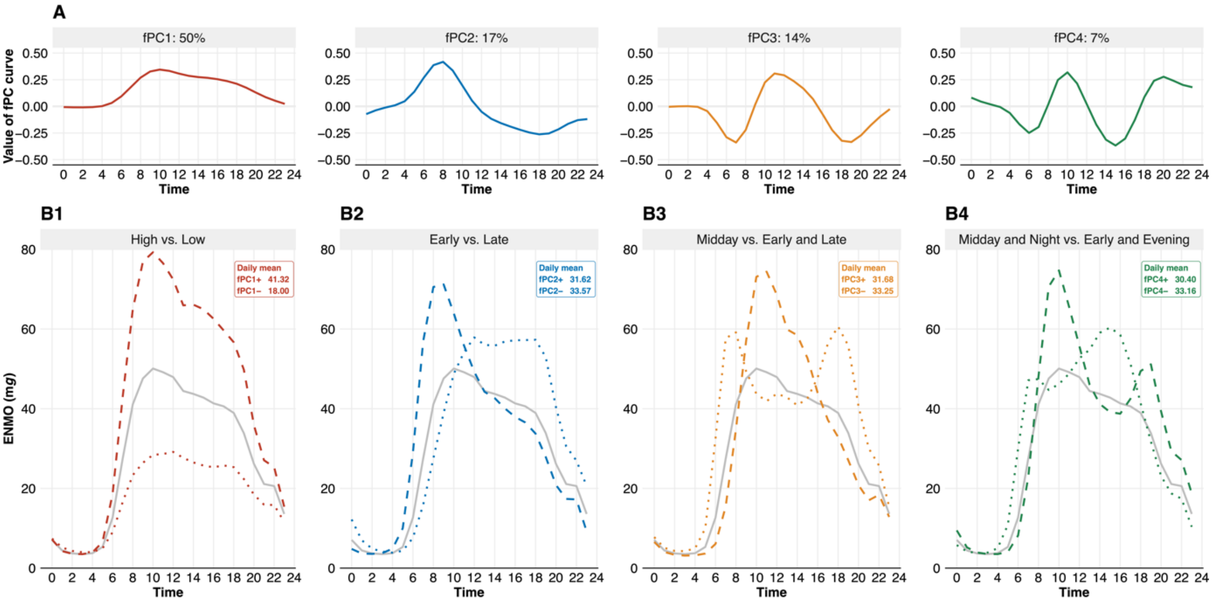


### S5C: Hazard ratios when using an Epanechnikov kernel

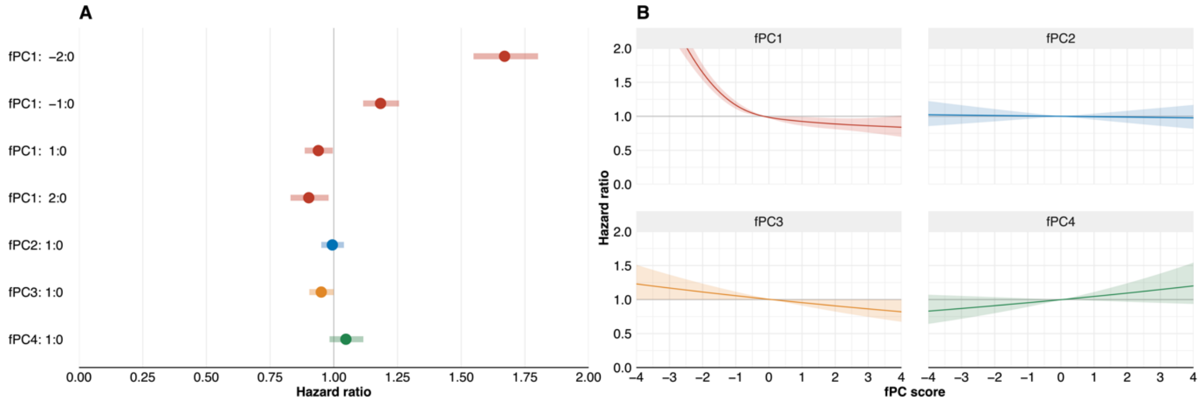

**S6: Cox models without deaths within 2 years after accelerometry and without prevalent CVD and/or diabetes**

| Component | Model with deaths within 2 years excluded HR (95% CI) | Model with prevalent CVD and/or diabetes excluded HR (95% CI) |
|---|---|---|
| fPC1 | | |
| *-2:0* | 1.61 (1.48–1.75) | 1.76 (1.59–1.96) |
| *-1:0* | 1.17 (1.10–1.26) | 1.22 (1.13–1.31) |
| *1:0* | 0.95 (0.88–1.01) | 0.93 (0.86–1.01) |
| *2:0* | 0.91 (0.83–1.00) | 0.91 (0.81–1.02) |
| *Overall p-value* | $6.29 \times 10^{-38}$ | $3.31 \times 10^{-35}$ |
| fPC2 | 0.96 (0.91–1.01) | 0.99 (0.93–1.06) |
| *Overall p-value* | 0.087 | 0.783 |
| fPC3 | 0.91 (0.86–0.97) | 0.92 (0.86–0.99) |
| *Overall p-value* | 0.005 | 0.025 |
| fPC4 | 1.08 (0.99–1.18) | 1.11 (1.00–1.23) |
| *Overall p-value* | 0.088 | 0.054 |

Preprint

### S7: Interaction of fPC1 and age groups

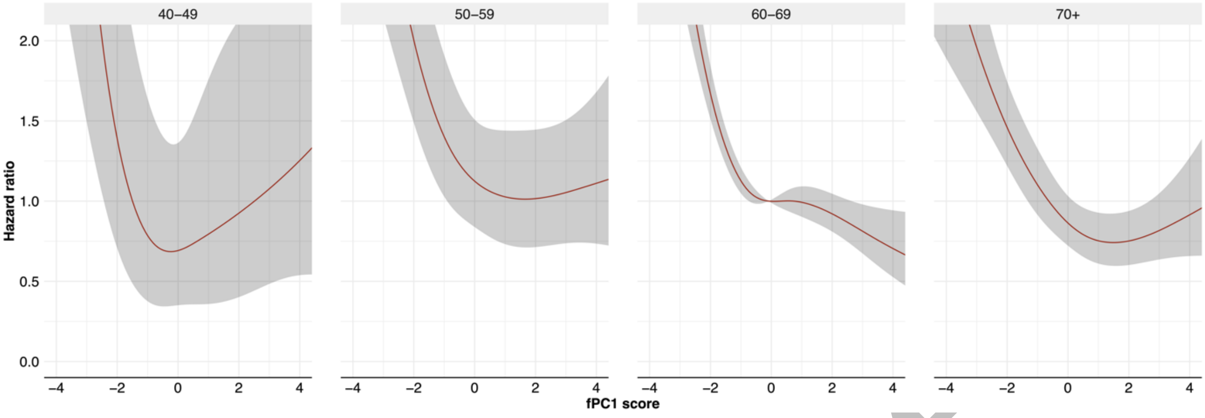


Preprint
