## Supplementary material for "Association between circadian physical activity patterns and mortality in the UK Biobank": STROBE checklist

STROBE Statement—Checklist of items that should be included in reports of ***cohort studies***

| | Item No | Recommendation |
|---|---|---|
| **Title and abstract** | 1 | (*a*) Indicate the study's design with a commonly used term in the title or the abstract<br>*The abstract describes the study design as "population-based prospective cohort study" and names the study (UK Biobank) used for the research*<br>(*b*) Provide in the abstract an informative and balanced summary of what was done and what was found<br>*The abstract describes methods and findings* |
| **Introduction** | | |
| Background/rationale | 2 | Explain the scientific background and rationale for the investigation being reported<br>*The background and rationale are described in the "Introduction" section* |
| Objectives | 3 | State specific objectives, including any prespecified hypotheses<br>*The objective of the study is described in the "Introduction" section (last sentence)* |
| **Methods** | | |
| Study design | 4 | Present key elements of study design early in the paper<br>*The study design is described in the section "Study population and data collection" (first subsection in the Methods section)* |
| Setting | 5 | Describe the setting, locations, and relevant dates, including periods of recruitment, exposure, follow-up, and data collection<br>*This information is described in the sections "Study population and data collection" and "Cohort follow-up and ascertainment of mortality cases"* |
| Participants | 6 | (*a*) Give the eligibility criteria, and the sources and methods of selection of participants. Describe methods of follow-up<br>*Eligibility criteria is described in the sections "Study population and data collection" and "Physical activity data" and Supplements S2 shows a Flow chart. Follow-up is described in the section "Cohort follow-up and ascertainment of mortality cases"*<br>(*b*) For matched studies, give matching criteria and number of exposed and unexposed<br>*Not applicable* |
| Variables | 7 | Clearly define all outcomes, exposures, predictors, potential confounders, and effect modifiers. Give diagnostic criteria, if applicable<br>*Exposures are described in the section "Physical activity data". Potential confounders are described in the section "Covariates". Outcomes are described in the section "Cohort follow-up and ascertainment of mortality cases"* |
| Data sources/ measurement | 8* | For each variable of interest, give sources of data and details of methods of assessment (measurement). Describe comparability of assessment methods if there is more than one group<br>*Exposures are described in the section "Physical activity data". Outcomes are described in the section "Cohort follow-up and ascertainment of mortality cases"* |
| Bias | 9 | Describe any efforts to address potential sources of bias<br>*Adjustment methods and sensitivity analyses to address potential sources of bias are described in the sections "Functional Principal Component Analysis (fPCA)" and „Statistical analysis"* |

Preprint

| | | |
|---|---|---|
| Study size | 10 | Explain how the study size was arrived at<br>*The study sample is described in the sections "Study population and data collection", "Physical activity data", "Statistical analysis", and in the flow chart (Supplement S2)* |
| Quantitative variables | 11 | Explain how quantitative variables were handled in the analyses. If applicable, describe which groupings were chosen and why<br>*Use of variables is described in the sections "FunctionalPprincipal ComponentAanalysis (fPCA)" and "Statistical analysis"* |
| Statistical methods | 12 | (*a*) Describe all statistical methods, including those used to control for confounding<br>*Statistical methods are discussed in the „Statistical analysis" section*<br>(*b*) Describe any methods used to examine subgroups and interactions<br>*Described in the section „Statistical analysis"*<br>(*c*) Explain how missing data were addressed<br>*Described in the section "Physical activity data" and „Statistical analysis"*<br>(*d*) If applicable, explain how loss to follow-up was addressed<br>*Not applicable*<br>(*e*) Describe any sensitivity analyses<br>*Described in the sections "Functional Principal Component Analysis (fPCA)" and „Statistical analysis"* |
| **Results** | | |
| Participants | 13* | (a) Report numbers of individuals at each stage of study—eg numbers potentially eligible, examined for eligibility, confirmed eligible, included in the study, completing follow-up, and analysed<br>*Described in the sections "Study population and data collection" and "Physical activity data" and shown in the flow chart (Supplement S2)*<br>(b) Give reasons for non-participation at each stage<br>*See flow chart (Supplement S2)*<br>(c) Consider use of a flow diagram<br>*See flow chart (Supplement S2)* |
| Descriptive data | 14* | (a) Give characteristics of study participants (eg demographic, clinical, social) and information on exposures and potential confounders<br>*Baseline characteristics are given in Tables 1 and Supplement S4*<br>(b) Indicate number of participants with missing data for each variable of interest<br>*Presented in flow chart in Supplement S2 and Table 1 and Supplement S4*<br>(c) Summarise follow-up time (eg, average and total amount)<br>*Presented in the first paragraph of the "Results" section* |
| Outcome data | 15* | Report numbers of outcome events or summary measures over time<br>*Number of cases are reported in the first paragraph of the "Results" section* |

Preprint

| | | |
|---|---|---|
| Main results | 16 | (*a*) Give unadjusted estimates and, if applicable, confounder-adjusted estimates and their precision (eg, 95% confidence interval). Make clear which confounders were adjusted for and why they were included<br>*Adjusted estimates are shown in Table 3 and Supplement S6 and are described in the "Results" section*<br>(*b*) Report category boundaries when continuous variables were categorized<br>*Exposures were not categorized*<br>(*c*) If relevant, consider translating estimates of relative risk into absolute risk for a meaningful time period<br>*Not applicable* |
| Other analyses | 17 | Report other analyses done—eg analyses of subgroups and interactions, and sensitivity analyses<br>*Sensitivity analyses are described in the section "Statistical analysis" and their results are reported in the "Results" section and in Supplements S5-7* |
| **Discussion** | | |
| Key results | 18 | Summarise key results with reference to study objectives<br>*Results are summarised in the first paragraph of the "Discussion" section* |
| Limitations | 19 | Discuss limitations of the study, taking into account sources of potential bias or imprecision. Discuss both direction and magnitude of any potential bias<br>*Study limitations are discussed in the "Discussion" section* |
| Interpretation | 20 | Give a cautious overall interpretation of results considering objectives, limitations, multiplicity of analyses, results from similar studies, and other relevant evidence<br>*Overall interpretation is given in the "Conclusion" section* |
| Generalisability | 21 | Discuss the generalisability (external validity) of the study results<br>*Generalisability is discussed in the "Discussion" section* |
| **Other information** | | |
| Funding | 22 | Give the source of funding and the role of the funders for the present study and, if applicable, for the original study on which the present article is based<br>*Stated in the "Funding/Support" section* |

*Give information separately for exposed and unexposed groups.

**Note:** An Explanation and Elaboration article discusses each checklist item and gives methodological background and published examples of transparent reporting. The STROBE checklist is best used in conjunction with this article (freely available on the Web sites of PLoS Medicine at http://www.plosmedicine.org/, Annals of Internal Medicine at http://www.annals.org/, and Epidemiology at http://www.epidem.com/). Information on the STROBE Initiative is available at http://www.strobe-statement.org.

Preprint
